# Genetic dissection of hippocampal sclerosis of ageing using magnetic resonance imaging surrogates

**DOI:** 10.1101/2025.03.10.25323307

**Authors:** Clàudia Olivé, Itziar de Rojas, Linda Zhang, Oscar Sotolongo-Grau, Inés Quintela, Pablo García-González, Raquel Puerta, Fernando García-Gutiérrez, Laura Montrreal, Maria Capdevila-Bayo, Andrea Miguel, Josep Blazquez-Folch, Miguel Calero, Alberto Rábano, Ana Belén Pastor, Teodoro del Ser, Miguel Medina, Ángel Carracedo, Alfredo Ramírez, Laura Molina-Porcel, Lluís Tàrraga, Amanda Cano, Sergi Valero, Marta Marquié, Pascual Sánchez-Juan, Mercè Boada, Bryan Strange, Maria Victoria Fernández, Agustín Ruiz

**Author notes:** These authors contributed equally to this work (Clàudia Olivé, Itziar de Rojas). . 57 C. de Marquès de Sentmenat, 08029 Barcelona, Spain (Maria Victoria Fernández).

## Abstract

**INTRODUCTION:** Hippocampal sclerosis of aging (HS-aging) is frequently present in individuals over 85 who die with dementia. Recent studies suggest that some loci associated with Alzheimer’s disease (AD) may be more related to HS-aging. We aimed to find AD-associated SNPs potentially related to HS-aging.

**METHODS:** We assessed the relation of the AD polygenic risk score (AD-PRS) with hippocampal subfield volumes assessed by magnetic resonance imaging (MRI) as HS-by-proxy in 1,130 non-demented participants. We analyzed 1,708 individuals to associate their AD-PRS (83 variants) with AD alongside HS-aging.

**RESULTS:** HS-by-proxy measures of fimbria and hippocampal body and head show association with AD-PRS, *SHARPIN*, *GRN* and *TNIP1*, also after replication. We replicated the stronger AD-PRS association with AD in the presence of HS-aging compared to AD alone.

**DISCUSSION:** Results show association between some AD-SNPs and HS-proxy, enriched in immune-brain axis pathways, differentiating HS-aging from AD. This insight aids in understanding their interrelationships and identifying specific therapeutic targets.

## 1. BACKGROUND

Hippocampal sclerosis of aging (HS-aging) is present in a considerable proportion of people over 85 years old who die with dementia [1], although magnetic resonance imaging (MRI) correlates of this disease are evident over a decade prior [2]. The main pathological features of HS-aging are severe neuronal loss and gliosis in specific regions of the hippocampus, causing cognitive and memory symptoms that appear to mimic Alzheimer’s disease (AD). Yet, HS-aging has usually less severe symptoms and longer evolution than AD [3] because it does not progress outside the hippocampus. HS-aging is also strongly associated with TDP-43 pathology and is encompassed in limbic-predominant age-related TDP-43 encephalopathy (LATE), which might also be an AD co-pathology [1,4]. Atrophy in hippocampal subfields detected by (MRI) has been recently reported to be a promising *in vivo* biomarker specific to HS-aging [5]. However, due to the similarity of their clinical profiles, and considering the current lack of biomarkers for HS-aging detection, this condition is often misdiagnosed as AD [1,5,6]. Further research is needed to dissect the genetic component and specific biomarkers for HS-aging, as well as its relationships with AD, LATE and other dementias.

The etiology of HS-aging is mostly unknown, but it is believed to involve several factors. HS-aging usually occurs in association with traumatic brain lesions (caused by events such as head injuries or seizures) and inflammation [7]. Furthermore, various studies have reported some evidence for genetic predisposition to HS-aging [8,9], indicating that the risk of HS-aging following a brain insult might depend partially upon genetic background [7].

Common diseases, such as AD or HS-aging, are complex traits which are rarely caused by the dysfunction of a single gene, but rather affected by the additive contribution of several genetic variants together [10]. Through genome-wide association studies (GWAS), it is possible to identify genetic variants significantly present in case individuals and map its polygenic architecture [10], and use them to construct polygenic risk scores (PRS) to identify individuals at risk, improve diagnostic tools and develop new drug targets [11].

Over the last decade, GWAS have grown significantly in sample size and in the number of traits under investigation. Recently, thanks to large international collaborative efforts [11,12], the genomic landscape of the AD risk alleles has doubled, with more than 80 AD-associated loci identified [12]. However, adding a larger sample size of clinically labeled individuals may result in the addition of cases with co-pathologies, and often misdiagnosis [6,13]. In fact, more than half of individuals with AD brain pathology are found to also have one or more other brain abnormalities that may cause dementia, such as vascular lesions, Lewy bodies, LATE, argyrophilic grains or HS-aging [14].

Hippocampal atrophy is observed in both healthy individuals and patients with mild cognitive impairment (MCI) as prodromal stage of AD dementia [15,16]. Thus, previous studies have associated the AD polygenic risk score (AD-PRS) with reduced volume in different hippocampal subfields in healthy individuals [17,18]. Nevertheless, some studies suggest that hippocampal atrophy before dementia onset manifests earlier in individuals with HS-aging compared to those with AD [19], supporting its role as an early biomarker for HS-aging [5,19,20]. Therefore, we hypothesize that certain variants included in the AD-PRS might be specifically associated with HS-aging.

In this study we aimed to assess the association of AD risk alleles with HS-aging using different hippocampal subfield volumes measured with MRI in non-demented participants. Since the genomic architecture has not been as extensively studied in HS-aging as in AD, we designed our analyses to detect which AD-related SNPs [12] might be more specifically associated with hippocampal atrophy patterns related to HS-aging. To achieve this, we examined both the individual and combined effects of AD-related SNPs included in the AD-PRS by Bellenguez et al. [12] and investigated their correlation with HS-aging.

## 2. METHODS

### 2.1. Participants

For this study we leveraged data from Ace Alzheimer Center Barcelona (ACE) [21], Alzheimer’s Disease Neuroimaging Initiative (ADNI, adni.loni.usc.edu, [22,23]) and the Vallecas Project (VP) [24]. We combined data from ACE and ADNI for our discovery analyses whereas VP was used for replication purposes. All samples for this study have available MRI, GWAS and clinical data. Participants from ACE (N=263) were MCI and non-demented collected through the BIOFACE project [25], Fundació ACE Healthy Brain Initiative (FACEHBI) [26], European Prevention of Alzheimer’s Dementia (EPAD) [27] and Models of Patient Engagement for Alzheimer’s Disease (MOPEAD) [28]. From ADNI we selected participants from either ADNI1 or ADNI2 who had a diagnosis of MCI (N = 665) or were non-demented (N = 465). For our replication analyses, we leveraged a cohort of non-demented individuals (N = 729) from the VP [24]. A summary of demographic and clinical information for the participants included in this study by project is shown in Table 1, and further information about each individual project is provided in the Supplementary Methods. All projects passed the ethics committee and written informed consent was obtained from all participants. Moreover, the EADB/ACE/BTN-Clinic-FRCB-IDIBAPS cohort (Supplementary Methods) was used for the replication of the association between AD-PRS and AD concomitant pathologies previously published by de Rojas et al. [11].

**Table 1.** Summary statistics. . Demographic and clinical data for the subjects included in the study by project.

| Projects | N | Females (%) | MCI (%) | CN (%) | Age (±SD) | Education (±SD) |
| --- | --- | --- | --- | --- | --- | --- |
| **Discovery** |  |  |  |  |  |  |
| ADNI-1 | 562 | 221(39.32) | 353(62.81) | 209(37.19) | 75.12(6.51) | 15.83(2.97) |
| ADNI-2 | 305 | 141(46.23) | 212(69.51) | 93(30.49) | 71.89(7.24) | 16.16(2.58) |
| BIOFACE | 56 | 37(66.07) | 56(100) | 00(00.00) | 61.46(3.48) | 9.77(3.05) |
| FACEHBI | 158 | 99(62.66) | 2(1.27) | 156(98.73) | 67.27(6.90) | 12.98(4.59) |
| EPAD | 29 | 15(51.72) | 23(79.31) | 6(20.69) | 70.68(5.82) | 9.79(3.48) |
| MOPEAD | 20 | 9(45.00) | 19(95.00) | 1(5.00) | 73.79(4.63) | 9.10(4.06) |
| Total | 1,130 | 522(46.19) | 665(58.84) | 465(41.15) | 72.34(7.55) | 14.95(3.75) |
| **Replication** |  |  |  |  |  |  |
| VP | 729 | 496(68.04) | 00(00.00) | 729(100) | 74.74(3.84) | 10.30(5.77) |
| Total | 1,859 | 1,018(54.76) | 665(35.77) | 1,194(64.22) | 73.28(6.47) | 13.13(5.17) |

\* N: number of samples; MCI: Mild cognitive impairment; CN: Controls; Age: average age at MRI; SD: Standard error.

**Table 2.** HS-aging and AD-PRS association. . Meta-analysis results of the hippocampal subfield volumes association with AD-PRS [13].

| Hippocampal subfield | OR | CI (95%) | P value |
| --- | --- | --- | --- |
| **Discovery (N = 1,128)** |  |  |  |
| Parasubiculum | 0.96 | 0.91-1.02 | $1.67 \times 10^{-01}$ |
| Hippocampal fissure | 1.02 | 0.96-1.08 | $5.16 \times 10^{-01}$ |
| Fimbria | 0.92 | 0.87-0.97 | **$2.13 \times 10^{-03}$** |
| Hippocampal BH | 0.91 | 0.87-0.96 | **$1.42 \times 10^{-04}$** |
| **Replication (N = 728)** |  |  |  |
| Fimbria | 0.95 | 0.89-1.02 | $1.76 \times 10^{-01}$ |
| Hippocampal BH | 0.94 | 0.88-1.00 | **$3.55 \times 10^{-02}$** |
| **Final (N = 1,856)** |  |  |  |
| Fimbria | 0.93 | 0.89-0.97 | **$1.12 \times 10^{-03}$** |
| Hippocampal BH | 0.92 | 0.89-0.96 | **$1.77 \times 10^{-05}$** |

\* Significant *P* values after Bonferroni correction in bold (*P* value < .0125; replication *P* value < .05).
† OR: Odds ratio; CI: Confidence interval; BH: Body and head.

### 2.2. Genomic data processing, QC and PRS

Samples from ACE and VP were genotyped as part of the Genome Research at Ace Alzheimer center (GR@ACE) and Dementia Genetics Spanish Consortium (DEGESCO) genetic initiatives [11,29]. Genotyping was conducted using the Axiom 815K Spanish biobank array (according to manufacturer’s instructions - Axiom™ 2.0 Assay Manual Workflow, Thermo Fisher) at the Spanish National Center for Genotyping (CeGEN, Santiago de Compostela, Spain). Details on genotyping and quality control procedures are provided elsewhere [29].

ADNI samples were genotyped with the Illumina Human 610-Quad BeadChip (Illumina, Inc., San Diego, CA, USA), for ADNI-1 and with the OmniExpress BeadChip for ADNI-GO/2 individuals (ADNI-2) [30,31].

Genomic data quality control (QC) was performed uniformly in three parallel batches for ADNI-1, ADNI-2 and ACE-VP. QC included removal of samples with low-quality genotyping, excess of heterozygosity or high missingness and variants with call rate below 95% or deviation from the Hardy–Weinberg equilibrium (*P* value < 1×10^-06^). Additionally, samples with discordant genetic sex annotation, family members (PI-HAT > 0.1875), or with a non-European ancestry (as per 1000 Genomes Project) were excluded from the analysis.

To maximize the AD-PRS coverage, imputed data was generated with the Trans-Omics for Precision Medicine (TOPMed) reference panel [32,33] on genome build GRCh38. Rare variants (MAF < 1%) and variants with low imputation quality (R^2^ < 0.3) were excluded.

A weighted individual PRS was calculated based on the 83 genome-wide significant (GWS, *P* value < 5E-08) variants reported by the European Alzheimer’s and Dementia Biobank (EADB) [12]. AD-PRS was generated by multiplying the genotype dosage of each risk allele for each variant by its respective weight and then summing across all variants. Due to its large effect, we excluded *APOE* variants (rs429358 and rs7412) from the PRS calculation as well as the *ABI3* locus (rs616338) because it was not properly imputed in the ADNI-2 dataset.

### 2.3. Structural MRI image acquisition

The image acquisition process was slightly different for each project of the ACE cohort included in this study. MRI scans from FACEHBI were performed on a 1.5T Siemens MAGNETOM Aera (Erlangen, Germany), while for for BIOFACE, EPAD and MOPEAD images were acquired with a Siemens MAGNETOM VIDA 3T scanner (Erlangen, Germany) using a 32-channel head coil at Clínica Corachan, Barcelona. Anatomical T1-weighted images were acquired using a rapid acquisition gradient-echo three-dimensional (3D) magnetization-prepared rapid gradient-echo (MPRAGE) sequence with different parameters for FACEHBI [26] (repetition time (TR) 2.200 ms, echo time (TE) 2.66 ms, inversion time (TI) 900 ms, slip angle 8°, field of view (FOV) 250 mm, slice thickness 1 mm, and isotropic voxel size 1 × 1 × 1 mm), BIOFACE [25] (TR 2.200ms, TE 2.23 ms, TI 968ms, 1.2 mm slice thickness, FOV 270 mm, and voxel measurement 1.1 × 1.1 × 1.2mm), MOPEAD [28] (TR 2.200 ms, TE 2.33 ms, TI 968ms, slip angle 8°, FOV 270 mm, slice thickness 1.2 mm, and isotropic voxel size 1.1 × 1.1 × 1.2 mm) and EPAD [27] (TR 2.300 ms, TE 2.93 ms, TI 900ms, slip angle 9°, FOV 270 mm, slice thickness 1.2 mm, and isotropic voxel size 1.1 × 1.1 × 1.2 mm). Axial T2-weighted, 3D isotropic fast fluid-attenuated inversion recovery (FLAIR) and axial T2*-weighted sequences were also acquired to detect significant vascular brain damage or microbleeds.

All the subjects with existing GWAS information were chosen from ADNI cohorts (http://adni.loni.usc.edu/). The closest MRI to the baseline was selected for each subject. All MRI T1-weighted images were downloaded in NIfTI format [34,35]. If available, more than one T1-weighted MRI for the same experiment was downloaded, in order to perform movement correction in the Freesurfer image processing pipeline (described below).

For the VP cohort, all T1-weighted images (3D fast spoiled gradient echo with inversion recovery preparation) were acquired using a 3T MRI (Signa HDxt GEHC, Waukesha, USA) with a phased array 8 channel head coil and the following parameters: repetition time (TR) 10 ms, echo time (TE) 4.5 ms, inversion time (TI) 600 ms, field of view (FOV) 240 mm, matrix 288×288 and slice thickness 1 mm, yielding 0.5×0.5×1 mm voxel size. All MRI scans were reported by a neuroradiologist.

### 2.4. Hippocampal subfields extraction

In order to extract the hippocampal subfield volumes, subjects from ACE and ADNI were processed the same way. First, cortical reconstruction and volumetric segmentation for MRI images was performed with the Freesurfer 7.2 image analysis suite, which is documented and freely available for download online (https://surfer.nmr.mgh.harvard.edu/). The technical details of these procedures are described in prior publications [36–38]. Briefly, this processing includes motion correction and averaging [39] of multiple volumetric T1-weighted images (when more than one is available), removal of non-brain tissue using a hybrid watershed/surface deformation procedure [40], automated Talairach transformation, segmentation of the subcortical white matter and deep gray matter volumetric structures (including hippocampus, amygdala, caudate, putamen, ventricles) [41,42] intensity normalization [43], tessellation of the gray matter white matter boundary, automated topology correction [44,45], and surface deformation following intensity gradients to optimally place the gray/white and gray/cerebrospinal fluid borders at the location where the greatest shift in intensity defines the transition to the other tissue class [36–38].

Then, the hippocampal subfields segmentation (HSF) was carried out using the hippocampal parcellation method included in Freesurfer 7.2. This is a tool that uses a probabilistic atlas built with ultra-high-resolution ex vivo MRI data (∼0.1 mm isotropic) to produce an automated segmentation of the hippocampal substructures and the nuclei of the amygdala. Individual subfield volumes for cornu ammonis (CA) were extracted out from results and grouped into a table. The following subfields of the hippocampal formation were used for the analyses in this study: CA1 body CA1 head, CA3 body, CA3 head, CA4 body, CA4 head, Granule Cell and Molecular Layer of the Dentate Gyrus (GC-ML-DG) body, GC-ML-DG head, hippocampus-amygdala-transition-area (HATA), whole hippocampal body and head (BH), hippocampal tail, whole hippocampus, fimbria, hippocampal fissure, molecular layer of the hippocampus body, molecular layer of the hippocampal head, parasubiculum, presubiculum body, presubiculum head, subiculum body and subiculum head. Also, estimated total intracranial volumes (TIV) were extracted, from prior Freesurfer analyses, in order to make further volumetric corrections.

For the VP cohort, automatic segmentation of the hippocampus was performed on each participant’s T1-weighted image using FreeSurfer v.6.0 (https://surfer.nmr.mgh.harvard.edu/). Technical details of the whole-brain segmentation methods have been described previously [41]. Hippocampal volumes were extracted using the hippocampal subfields module in FreeSurfer 6.0 [46], and segmentations for all participants were visually inspected for accuracy.

### 2.5. Association of HS-proxy subfields with AD-PRS

Hippocampal subfield volumes and PRS data were standardized by project using the scale function in R (Supplementary Figure 1) and independent t-tests were conducted to assess differences between PRS means in individuals classified by diagnosis and *APOE* ɛ4 carriers. For hippocampal subfield volumes, outliers differing ±3 SD from the mean were removed. As has been frequently applied to MRI data analysis [47–50], we conducted hierarchical agglomerative clustering to group highly correlated hippocampal subfields. We used the average agglomeration method on the Euclidean distance matrix of hippocampal subfield volumes (Supplementary Figure 2 & 3). After dimension reduction of these variables, HS-by-proxy phenotype was based on the volumes of the following four hippocampal subfields: hippocampal fissure, parasubiculum, fimbria and whole hippocampal BH. All analyses were performed using R version 4.1.2 software.

Association of PRS with HS-by-proxy was assessed using linear regression for each project. HS-by-proxy and PRS were set as dependent and independent variables respectively, with sex, age, age^2^, years of education, diagnosis (when necessary), TIV and first ten genetic principal components (PC1-10) as covariates in the following regression model:

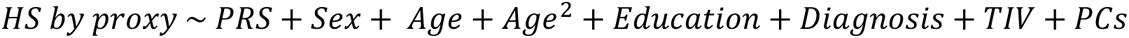

Pearson’s correlation analysis was conducted to study the association between each SNP included in the PRS calculation and HS-by-proxy phenotype previously found to be significantly associated with PRS. SNPs presenting higher and significant correlations (*P* value < .05 and Pearson’s r > .05) were later employed as independent variants for the estimation of linear regression models adjusted by the same covariates as before:

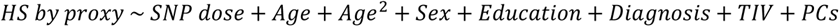

Results for both regression models were then combined across projects with a fixed-effect meta-analysis (I^2^ < 75%) for each hippocampal subfield representing HS-by-proxy phenotype, using the inverse variance weighted approach implemented with the *meta* package in R [51].

### 2.6. Co-regulatory network analysis

To identify co-regulatory networks of genes associated with HS-aging, we used GeneFriends [52], a bioinformatics pipeline previously reported by our team [53]. These lists of genes were based on the Sequence Read Archive (SRA) [54] and only genes highly co-expressed (co-expression value > 0.5) in neuron and/or brain tissue were included in further analyses. Next, WebGestalt [55] was used to identify potential enrichments in the previously identified co-regulated gene lists. We used over-representation enrichment analysis (ORA) [56] of Kyoto Encyclopedia of Genes and Genomes (KEGG) pathways in humans using protein-coding regions of the genome as reference set. Those lists of co-expressed genes showing significantly enriched KEGG pathways were further analyzed using STRING v11.5 [57] in order to find known and predicted protein-protein interactions.

## 3. RESULTS

### 3.1. AD-PRS and HS-by-proxy phenotype

In the meta-analysis presented here, we included data from six different projects, which consisted of 1,130 non-demented participants with MRI and GWAS data. Their demographic composition was as follows: 46.2% were female, 58.9% had MCI and 41.1% were non-demented (Table 1). The average age of the participants was 72.3 (±3.84) years. Additionally, an AD-PRS was calculated for these participants, which ranged from -3.60 to 3.73 on a scaled min-max basis (Supplementary Figure 4).

We found a study-wise significant association between the AD-PRS and hippocampal BH (OR [95% CI] = 0.91 [0.87–0.96]; *P* value = .000142) and fimbria (OR [95% CI] = 0.92 [0.87–0.97]; *P* value = .00213) as HS-by-proxy phenotypes after Bonferroni correction (*P* value < .0125).

Next, the Vallecas replication cohort (N = 728 healthy controls, 68.1% females and mean age 74.7 years with an AD-PRS range scaled _min-max_ from -3.11 to 2.96) confirmed a significant and independent association between AD-PRS and hippocampal BH (OR [95% CI] = 0.94 [0.88–1.00]; *P* value = .036) validating our discovery finding.

Combining all three cohorts in an extended meta-analysis (N = 1,856) showed the same effect size and improved the statistical significance of the association between the AD-PRS and the HS-by-proxy phenotypes (hippocampal BH, OR [95% CI] = 0.92 [0.89– 0.96]; *P* value = 1.77×10^−05^; fimbria, OR [95% CI] = 0.93 [0.89–0.97]; *P* value = .00112; Figure 1) suggesting that non-demented participants with higher genetic burden of AD have less volume in these hippocampal subfields. All subsequent values reported here correspond to results of analyses of all cohorts combined. The impact of covariates in each model is presented in Supplementary Table 1 for hippocampal BH and Supplementary Table 2 for fimbria.

**Figure 1.**
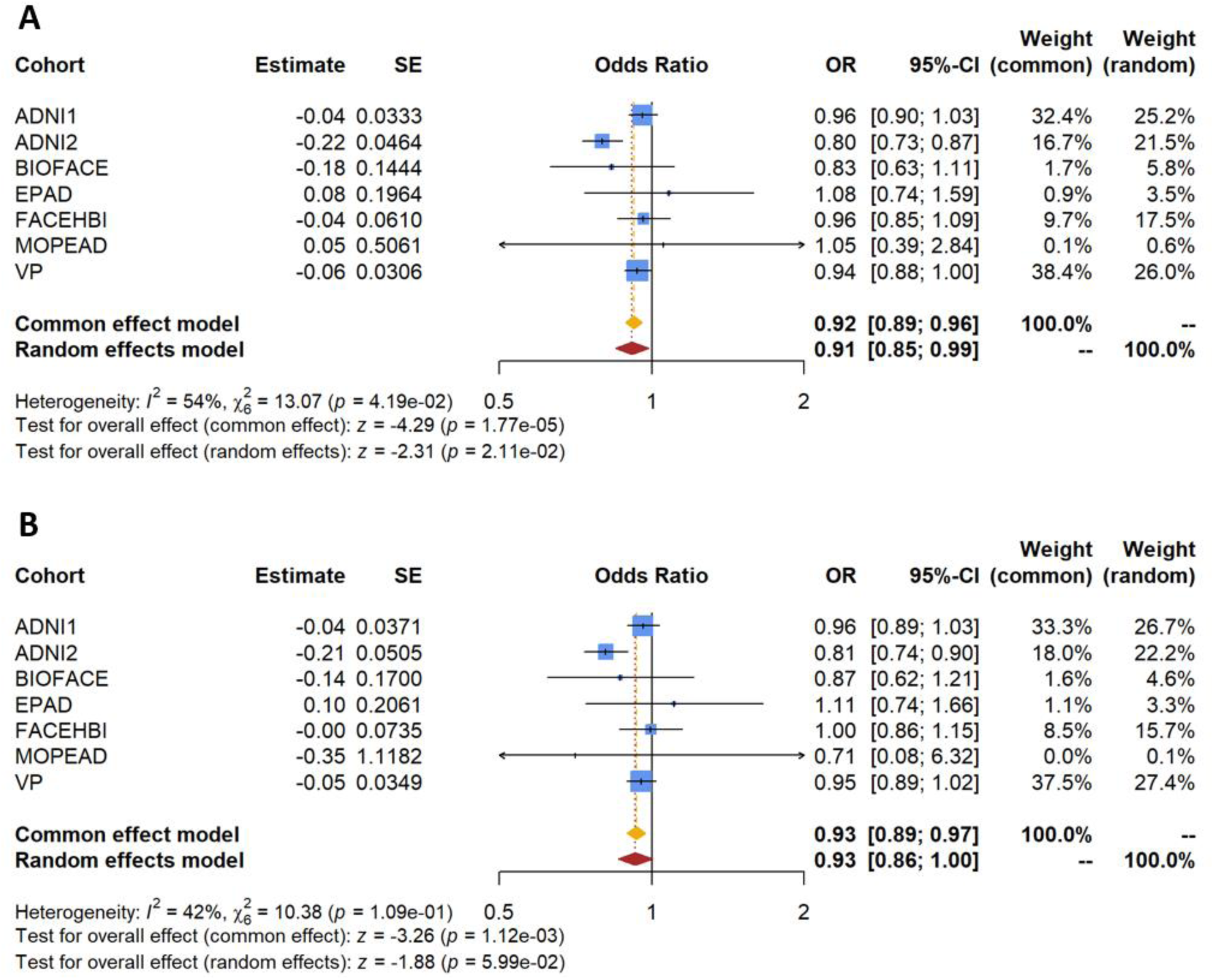
Meta-analysis for the association between AD-PRS and HS-by-proxy phenotypes. Forest plots for (**A**) hippocampal BH and (**B**) fimbria, including patients from discovery (ACE and ADNI) and replication (VP) cohorts.

### 3.2. AD SNPs and HS-by-proxy phenotype

For those HS-by-proxy phenotypes for which we found a significant association with AD-PRS (hippocampal BH and fimbria) we tested for correlation with the AD loci reported by Bellenguez et al. [12] composing the AD-PRS. The purpose of this analysis is to reduce the dimensionality of gene data associated with AD for a more specific assessment of its relation with HS-aging (Table 3). Variants in *SHARPIN* (rs34173062), *GRN* (rs5848) and *TNIP1* (rs871269) loci were found to be significantly associated with hippocampal BH (rs34173062, OR [95% CI] = 0.84 [0.77–0.92], *P* value = 1.51×10^−04^; rs871269, OR [95% CI] = 0.90 [0.85–0.96], *P* value = 3.57×10^−04^) and/or fimbria (rs34173062, OR [95% CI] = 0.81 [0.73–0.90], *P* value = 6.63×10^−05^; rs5848, OR[95% CI] = 0.89 [0.83–0.95], *P* value = 2.84×10^−04^; Figure 3) after Bonferroni correction (*P* value < 6.02×10^−04^) in the meta-analysis. Based on the correlation of AD-PRS loci with HS-by-proxy, we found three clusters of AD loci showing an unequal association profile to HS-aging which supports the existence of a loci set more related to specific hippocampal pathobiology (Figure 2). The SNPs in cluster A (blue cluster) show an overall negative correlation with our HS-by-proxy phenotype, meaning that they are generally correlated with smaller fimbria and hippocampal BH volumes, thus potentially specific to tissue atrophy. What is more, all SNPs found significantly associated with HS-by-proxy (*SHARPIN*, *GRN*, *TNIP1*) gather together in cluster A. SNPs in cluster B and C were not significantly associated with our HS-by-proxy phenotype, with an overall positive correlation among the SNPs in these clusters and hippocampal volume. This suggests a tendency toward greater hippocampal volumes for carriers of these cluster B and C variants.

**Figure 2.**
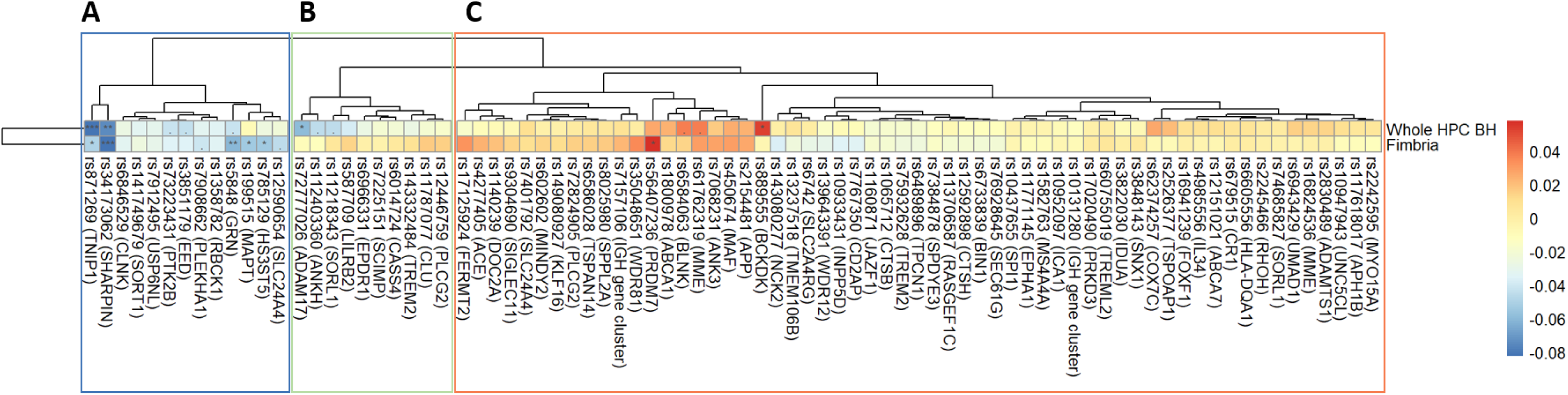
Correlations between HS-by-proxy phenotypes (Hippocampal BH and Fimbria) and AD SNPs included in the PRS in all study cohorts. (**A**) Framed in blue, cluster with a predominant negative Pearson’s correlation with HS-by-proxy phenotype represented in blue tones; (**B**) framed in green, cluster with a mixed positive (red) and negative (blue) Pearson’s correlation with HS-by-proxy phenotype; (**C**) cluster with a predominant positive correlation with HS-by-proxy phenotype represented in orange and red tones. All SNPs are represented by their risk allele for AD found by Bellenguez et al. [12]. HPC BH: Hippocampal body and head. Sig. level = 0.001***, 0.01**, 0.05*.

**Figure 3.**
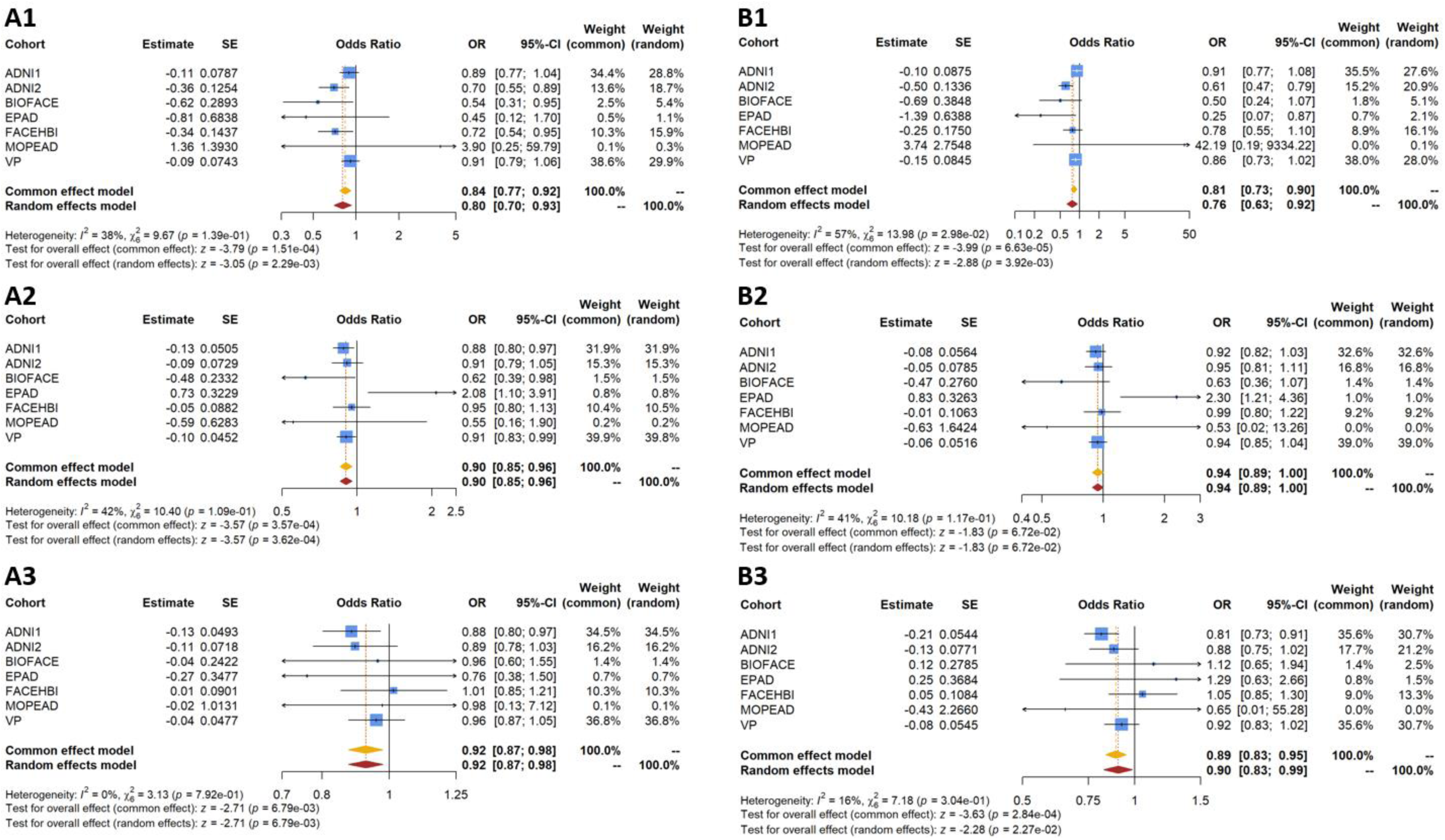
Meta-analysis for the association between the significative AD loci and HS-by-proxy phenotypes. Forest plots for *SHARPIN* (**1**), *TNIP1* (**2**) and *GRN* (**3**) loci with hippocampal BH (**A**) and fimbria (**B**) in all study cohorts.

**Table 3.** AD SNPs and HS-aging association. . Meta-analysis results of the HS-by-proxy association with the selected SNPs in the AD-PRS [13]. SNPs presenting higher correlations with HS-by-proxy phenotypes (*P* value < .05 and Pearson’s r > .05).

| Loci | SNP | OR | CI (95%) | $P$ value |
| --- | --- | --- | --- | --- |
| **Hippocampal BH** |  |  |  |  |
| *ADAM17* | rs72777026 | 0.90 | 0.84-0.97 | .0049 |
| *BCKDK* | rs889555 | 1.07 | 1.00-1.13 | .03 |
| *GRN* | rs5848 | 0.92 | 0.87-0.98 | .0068 |
| *HS3ST5* | rs785129 | 1.01 | 0.96-1.07 | .07 |
| *MAPT* | rs199515 | 0.92 | 0.87-0.98 | .01 |
| *PRDM7* | rs56407236 | 1.11 | 0.99-1.23 | .06 |
| *SHARPIN* | rs34173062 | 0.84 | 0.77-0.92 | **$1.51 \times 10^{-04}$** |
| *TNIP1* | rs871269 | 0.90 | 0.85-0.96 | **$3.57 \times 10^{-04}$** |
| **Fimbria** |  |  |  |  |
| *ADAM17* | rs72777026 | 0.95 | 0.88-1.03 | .25 |
| *BCKDK* | rs889555 | 1.00 | 0.93-1.07 | .99 |
| *GRN* | rs5848 | 0.89 | 0.83-0.95 | **$2.84 \times 10^{-04}$** |
| *HS3ST5* | rs785129 | 1.00 | 0.94-1.06 | .89 |
| *MAPT* | rs199515 | 0.89 | 0.83-0.96 | .0013 |
| *PRDM7* | rs56407236 | 1.19 | 1.05-1.34 | .0049 |
| *SHARPIN* | rs34173062 | 0.81 | 0.73-0.90 | **$6.63 \times 10^{-05}$** |
| *TNIP1* | rs871269 | 0.94 | 0.89-1.00 | .07 |

\* Significant $P$ values after Bonferroni correction in bold ( $P$ value $< 6.02 \times 10^{-04}$ ).
† OR: Odds ratio; CI: Confidence interval; BH: Body and head.

### 3.3. The LUBAC complex as represented pathway in the AD SNPs associated with HS-by-proxy phenotype

Using the STRING software, we found that cluster A (Figure 2; blue) which includes *SHARPIN, GRN* and *TNIP1*, was significantly enriched in the linear ubiquitin chain assembly complex (LUBAC) cellular component (Gene Ontology (GO) Term: GO:0071797) responsible for producing linear polyubiquitin chains and regulating the NF-κB pathway [58], which plays a critical role in inflammatory and immune responses. The biological processes enriched by this cluster are: protein linear polyubiquitination (GO:0097039) and the negative regulation of biological process (GO:0048519). Next, the top three significantly enriched biological process in cluster B (Figure 2B) were the regulation of neurofibrillary tangle assembly (GO:1902996), microglial cell proliferation (GO:0061518) and positive regulation of dendritic cell cytokine production (GO:0002732). In cluster C, the top three significantly enriched biological processes were the positive regulation of engulfment of apoptotic cell (GO:1901076), neuropeptide catabolic process (GO:0010813) and negative regulation of aspartic-type endopeptidase activity involved in amyloid precursor protein catabolic process (GO:1902960). To point out, the B and C clusters are both significantly enriched in their top ten pathways by the positive regulation of immune system process (GO:0002684). Another 23 biological processes are enriched by loci within both clusters B and C, with some of them being related to amyloid β (Aβ) formation or clearance. However, none of the pathways enriched for genes in cluster A are also enriched for loci in cluster B or C. Summaries of the biological processes enriched by each cluster are reported on Supplementary Table 3, 4 & 5.

Regarding disease-gene associations found using STRING, genes in clusters B and C, which are associated with higher hippocampal subfield volumes, showed a significant enrichment for AD (DOID:10652; B cluster FDR = 5.7×10^−03^; C cluster FDR = 8.8×10^−04^). In contrast to the cluster A, which includes genes correlated with smaller hippocampal subfield volumes and is only significantly enriched in nominal aphasia (DOID:4541; FDR = 1.84×10^−02^).

### 3.4. Network of genes associated with SNPs related to HS-by-proxy phenotype

Looking for a gene-network and pathways enriched for the genes associated with hippocampal BH and fimbria, we extracted the genes co-expressed with *SHARPIN, GRN* and *TNIP1* in brain tissue and neurons using Genefriends. For this purpose, only transcripts displaying a high expression correlation between them were selected (Pearson’s r > 0.5). We detected a 3.57% overlap among genes co-expressed with *SHARPIN, GRN* and *TNIP1* which were further tested in a pathway enrichment analysis (Supplementary Figure 5).

Nine significantly enriched KEGG pathways were identified by the enrichment analysis with WebGestalt and 10 pathways (FDR ≤ 0.05) with STRING in brain (Supplementary Table 6-7), No significant pathways were detected in neurons (Supplementary Table 8). Four of the most significantly enriched pathways concordant between the STRING and the WebGestalt strategies could be connected to biological mechanisms of HS-aging: protein processing in endoplasmic reticulum (hsa04141), other glycan degradation (hsa00511), B cell receptor signaling pathway (hsa04662) and lysosome (hsa04142).

## 4. DISCUSSION

Characterization of the genetic landscape of complex diseases provides a unique opportunity for a better understanding of their associated physiopathological processes. Although GWAS analyses have significantly helped improve our current understanding of AD genetic architecture, the massive sample sizes required for this kind of studies comes with the increased risk of adding higher rates of individuals with co-pathologies or doubtful diagnoses in these analyses. In fact, more than 50% of the people with dementia studied at U.S. AD Research Centers have evidence of more than one cause of dementia [59].

Our recent study of AD in a Spanish histopathological cohort [11], suggested a stronger association of the AD-PRS with AD-mixed pathologies than with AD alone. Updating the AD-PRS with the 83 SNPs from Bellenguez et al. [12] reveals an increase of the effect of this trend (Figure 4). These results indicate that the AD association of some AD-PRS variants might be due to their relation with other pathologies, like HS-aging.

**Figure 4.**
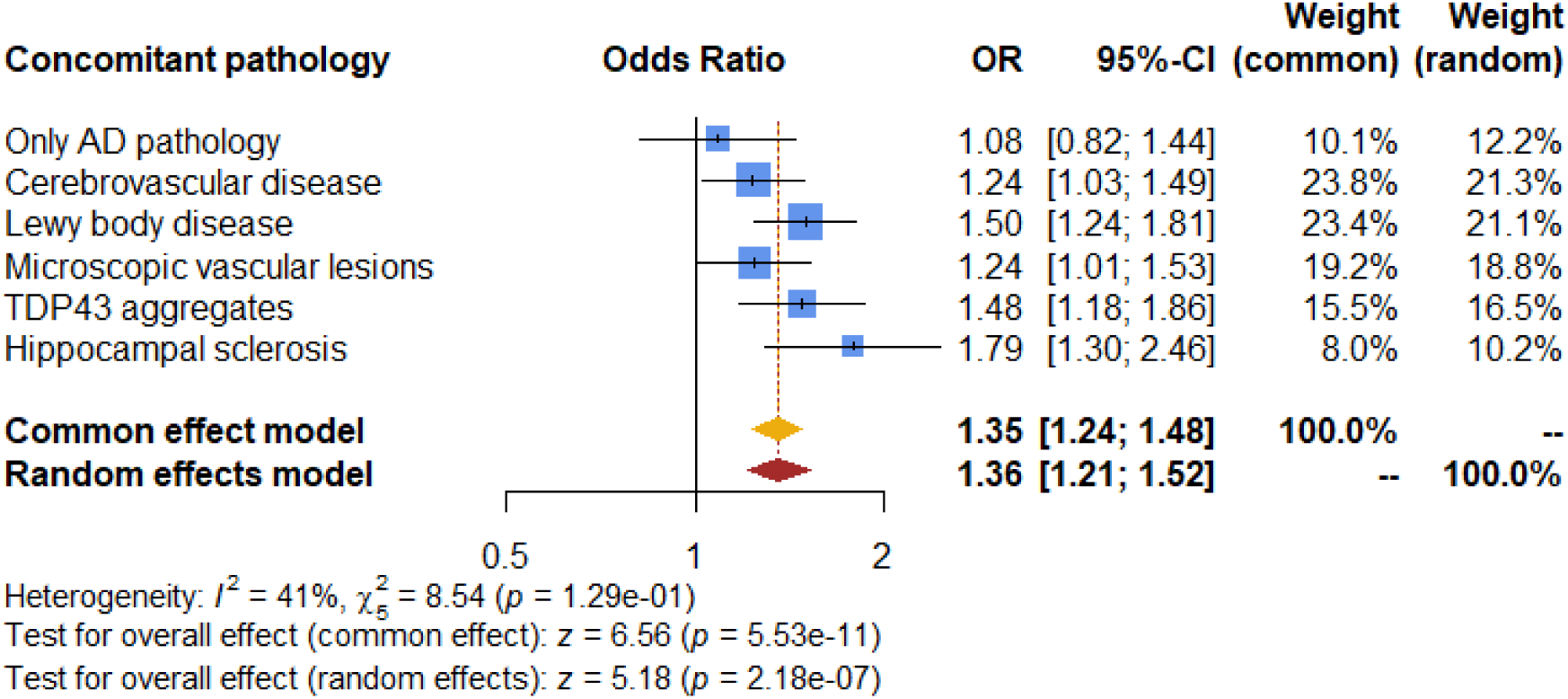
Meta-analysis for the association between the AD-PRS (based on Bellenguez et al. 2022) and AD in the presence of concomitant brain pathologies. Forest plots including data from EADB/ACE/BTN-Clinic-FRCB-IDIBAPS datasets from de Rojas et al. [11]. Data presented as OR per 1-SD increase in PRS (95% CI).

Our hypothesis arose from our previous findings [11] where AD-PRS was found to be more strongly associated with mixed pathologies (AD+HS-aging) than with pure AD pathology. We hypothesized that some SNPs associated with AD in large GWAS might be more specifically associated to HS-aging hippocampal atrophy patterns than AD. For this purpose, we used the AD-related SNPs extracted from the recently published meta-GWAS for AD risk [12] to assess their association with hippocampal subfield volumes as HS-by-proxy phenotype in individuals without dementia, since previous studies suggest that hippocampal atrophy seen in HS-aging begins early prior to dementia compared to AD and support its use as a biomarker for HS-aging [19,20].

In line with previous research that associate AD-PRS with hippocampal atrophy in non-demented individuals [17,18,60], the main findings of this study agree with our hypothesis: higher values of AD-PRS are associated not only with AD but more specifically with HS-by-proxy represented by small hippocampal subfields volumes in fimbria and hippocampus BH regions in individuals without clinical AD dementia. This is also in keeping with the recent demonstration [61] that *in vivo* MRI correlates of HS-aging, derived from postmortem histopathology, are primarily reductions in grey matter in anterior hippocampus (i.e., head extending into body). Regarding the loss of volume of with matter (i.e., the fimbria), we speculate that this occurs secondary to degeneration of the CA1 and subiculum. Moreover, our results show two new variants that are part of the AD-PRS but might be more specific to HS-aging (rs34173062 and rs871269). These variants drive the effect of the association between HS-aging and AD-PRS together with rs5848 in *GRN*, for which we replicate the association with HS-aging.

Previous HS-aging studies have consistently identified four loci (*GRN*, *TMEM106B*, *ABCC9*, and *KCNMB2*) [62]. Specifically, variant rs5848 in *GRN* has been associated with HS-aging [9,63]. *GRN* (Granulin Precursor) is a gene that encodes for granulins, a family of glycosylated peptides. Glycosylation happens in the endoplasmic reticulum (ER) and disruptions on the protein processing in the ER (KEGG pathway has04141, enriched by genes commonly co-expressed with *SHARPIN, GRN* and *TNIP1*) have been associated with neurodegenerative diseases. After being attached to proteins in the ER, glycans are degraded (KEGG pathway hsa00511) in the lysosome (KEGG pathway hsa04142) by autophagy, both pathways enriched by genes co-expressed with *SHARPIN* and *TNIP1*. Autophagy eliminates inflammatory triggers (i.e., cytokines) and regulates the organelle function in immune cells (i.e., B cells). The B cell receptor signaling pathway (KEGG pathway has04662), involved in inflammation [64], is enriched in our results by genes co-expressed with the loci linked with HS-aging.

Consistent with our findings, variant rs34173062 in *SHARPIN* has been suggested to be a genetic modifier of neuroanatomical variation in the limbic system through a GWAS of imaging that used a much larger sample size (N = 8,428) of younger individuals (age 49-69) [65]. Additionally, a rare variant in the *SHARPIN* gene (rs77359862), which is in linkage disequilibrium with the variant examined in this study (rs34173062, R^2^ = 0.003, D’= 1, and Minor Allele Frequency (MAF) = 3.5×10^−04^) and located 4,325 base pairs away, has been previously identified to have a genome-wide significant association with MRI traits in a Korean cohort [66]. *SHARPIN* (SHANK Associated RH Domain Interactor) is a gene coding for a postsynaptic density protein of excitatory synapses which is part of the NF-κB-activating the LUBAC complex in the nervous system[66]. NF-κB induces the expression of various pro-inflammatory genes, including those encoding cytokines and chemokines, and participates in inflammasome regulation [67]. Further supporting our results, variant rs871269 in *TNIP1* has been formerly associated with HS-aging pathology in 2,831 individuals with European ancestry [68]. *TNIP1* (TNFAIP3 Interacting Protein 1) is also a gene implicated in NF-κB activation and NF-κB-dependent gene expression involved in the anti-inflammatory response. Functionally, TNIP1 protein is an inflammation modulatory protein that exerts its influence by regulating nuclear factor κB activation [69].

All three variants in *SHARPIN*, *GRN* and *TNIP1* disclosed in this study, cluster with SNPs showing a negative correlation with hippocampal subfield volumes representing HS-by-proxy (Figure 2A). In addition, the only pathway enriched for this cluster, the LUBAC complex pathway, is associated with neurodegeneration via inflammatory pathways but neither with Aβ nor with Tau pathology. This, implies that these loci are potentially more specific to brain atrophy in general or hippocampal pathology in particular than to other AD pathological hallmarks. It is known that ubiquitination by the LUBAC complex is a key checkpoint in death receptor signaling [70]. Moreover, recent investigations demonstrate the impact of LUBAC-mediated linear polyubiquitination on the aggregation of disease associated proteins linked to various neurodegenerative diseases, such as TDP-43 proteinpathy which has been seen to improve after LUBAC inhibition [71]. Moreover, linear polyubiquitination by LUBAC complex leads the abnormal TDP-43 aggregates to autophagic proteolysis, via failed protein degradation system and subsequent NF-κB activation [72]. Given that *SHARPIN* is part of LUBAC and *TNIP1* is implicated in NF-κB activation, we speculate that these loci might be more prominently associated with TDP-43 and HS pathologies.

A previous study [73] adds evidence to the relation of these variants with the immune-mediated component of hippocampal atrophy. This study reported a pathway enrichment analysis of AD-loci by Bellenguez et al. [12] to the clusters obtained as a result of associating these AD SNPs with levels of Aβ42 and phosphorylated Tau (pTau) in cerebrospinal fluid (CSF). Variants in *GRN* and *TNIP1* cluster together separated from the variant located in *SHARPIN*. No GO-terms were enriched by the SNPs that cluster with *SHARPIN,* but *SHARPIN* alone shows a significant association with pTau in CSF [73], which may be related to pathologies other than AD. A total of eight GO-terms were enriched by *GRN* and *TNIP1* cluster. The common denominator of the names of the pathways enriched by this cluster is “immune”, of which *GRN* is one of the most frequents contributor. Aside from *GRN*, a high number of genes included in the “immune” cluster have been related to dementia types other than AD [73]. This aligns with the results presented in this study, which further confirm that these loci may be more closely related to dysfunction in the immune response involved in multiple neurodegenerative diseases rather than specifically to AD pathology [74].

The concept of the immune-brain axis has gained attention, highlighting bidirectional communication between the immune system and the central nervous system [75]. Since both age and neurodegeneration are associated with inflammatory processes, we speculate that in HS-aging, inflammatory responses in the hippocampus may influence autophagy events [74,75].

A limitation of our study is the heterogeneity of the MRI data due to varying acquisition parameters and Freesurfer versions across centers. Moreover, despite combining cohorts, our sample size remains limited, and a larger one is needed to increase statistical power and validate the present findings. A longitudinal follow-up of the present study to identify patients developing HS-aging, or future validations in HS-aging diagnosed or autopsy cohorts, is required to confirm our results. Nevertheless, this study yielded significant results and include independent and consistent replication results supporting our hypothesis.

In conclusion, AD-PRS and some AD-variants showed correlation with a proxy for HS-aging. By studying mixed dementia cases and HS-by-proxy, we identified HS-predominant genetic signals that might be instrumental for HS-aging risk detection. Specifically, variants in *SHARPIN, GRN* and *TNIP1* might be more related to HS-aging than to AD. Our study highlights the importance of precise phenotyping in genetic studies to generate disease-specific PRS. Dissecting the molecular pathways, cell types, and brain regions associated with each AD locus is crucial to translate genetic observations into clinical benefits.

## Data Availability

All data produced in the present study are available upon reasonable request to the authors.

## ACKNOWLEDGEMENTS

CO and IdR contributed to data acquisition, analysis, interpreted the data and co-wrote the manuscript. OSG and LZ contributed to MRI data analysis. ARu, MVF and IdR designed, conceptualized, supervised the study and interpreted the data. LT, SV, MMa, PSJ, MB, BS, MVF and ARu contributed to the critical revision of the paper. Data generation, sample contribution: CO, IdR, LZ, OSG, IQ, PGG, RP, FGG, LM, MCB, ACan, AM, JBF, MC, ARa, MTA, ABP, TdS, MMe and ACar. All authors critically revised the manuscript for important intellectual content and approved the final manuscript. We would like to extend our gratitude to participants and their families for their contribution of time and samples to this study. We are indebted to the Biobank-Hospital Clinic-FRCB-IDIBAPS for samples and data procurement. The present work has been performed as part of the doctoral program of C. Olivé at the Universitat de Barcelona (Barcelona, Spain).

## CONSENT STATEMENT

All human subjects provided informed consent.

## SUPPLEMENTARY FIGURES

**Supplementary Figure 1.** Distribution of the scaled HS-by-proxy phenotypic variables by project and clinical status. HPC (Hippocampal), BH (body and head), MCI (Mild Cognitive Impairment).

**Supplementary Figure 2.** Hippocampal subregions correlations. HP (Hippocampus), BH (body and head), HPC (Hippocampal), HATA (hippocampus-amygdala-transition-area), GC-ML-DG (Granule Cell and Molecular Layer of the Dentate Gyrus), CA (Cornu ammonis).

**Supplementary Figure 3.** Cluster dendrogram for hippocampal subfields. HP (Hippocampus), BH (body and head), HPC (Hippocampal), HATA (hippocampus-amygdala-transition-area), GC-ML-DG (Granule Cell and Molecular Layer of the Dentate Gyrus), CA (Cornu ammonis).

**Supplementary Figure 4.** Distribution of AD-PRS values in mild cognitive impairment (MCI) compared with control individuals (**A**). In *APOE* ɛ4 carriers (**B**) and *APOE* ɛ4 non-carriers (**C**).

**Supplementary Figure 5.** Venn diagram of the co-expressed genes between *SHARPIN, GRN* and *TNIP1* in brain (**A**) and neuron (**B**).

