## Supplementary Figures for "Genetic dissection of hippocampal sclerosis of ageing using magnetic resonance imaging surrogates"

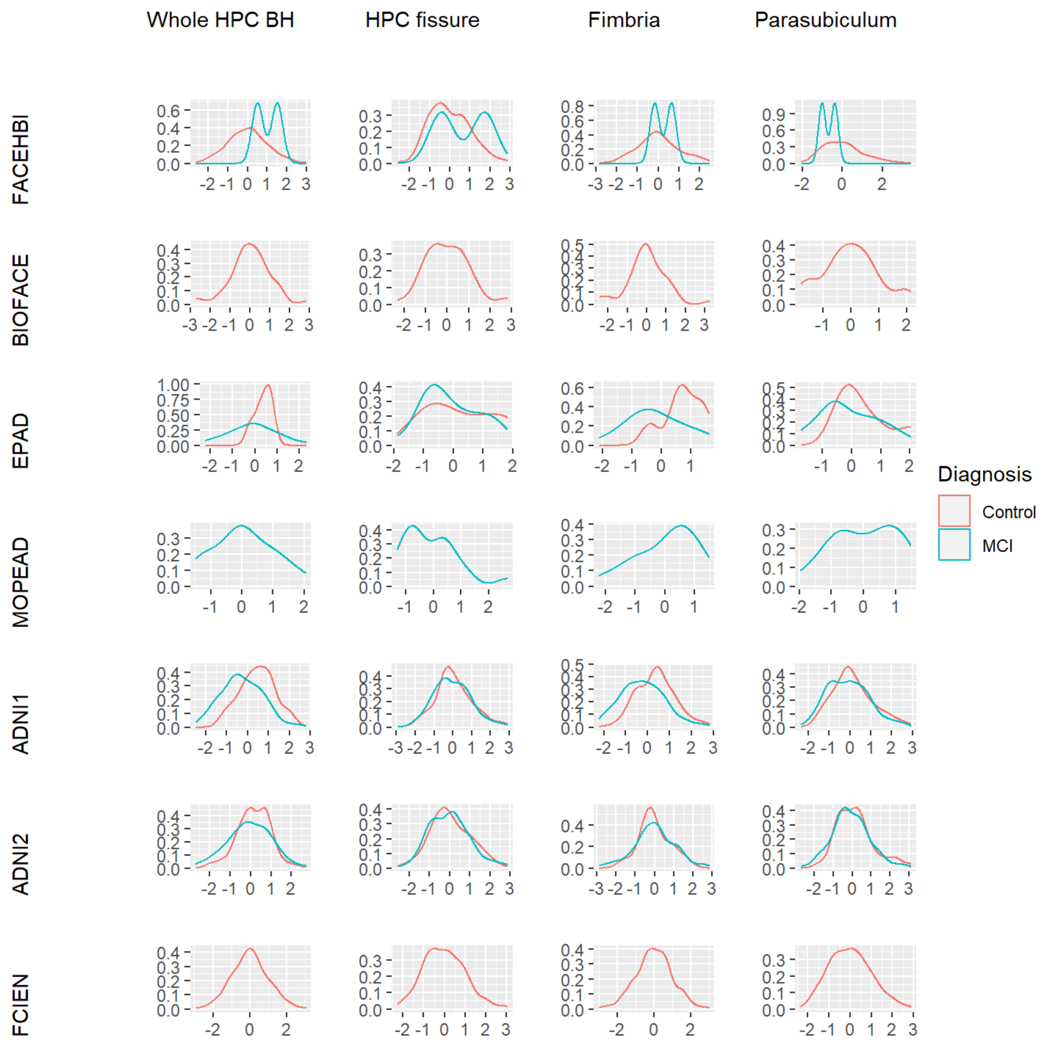


**Supplementary Figure 1**. Distribution of the scaled HS-by-proxy phenotypic variables by project and clinical status. HPC (Hippocampal), BH (body and head), MCI (Mild Cognitive Impairment).

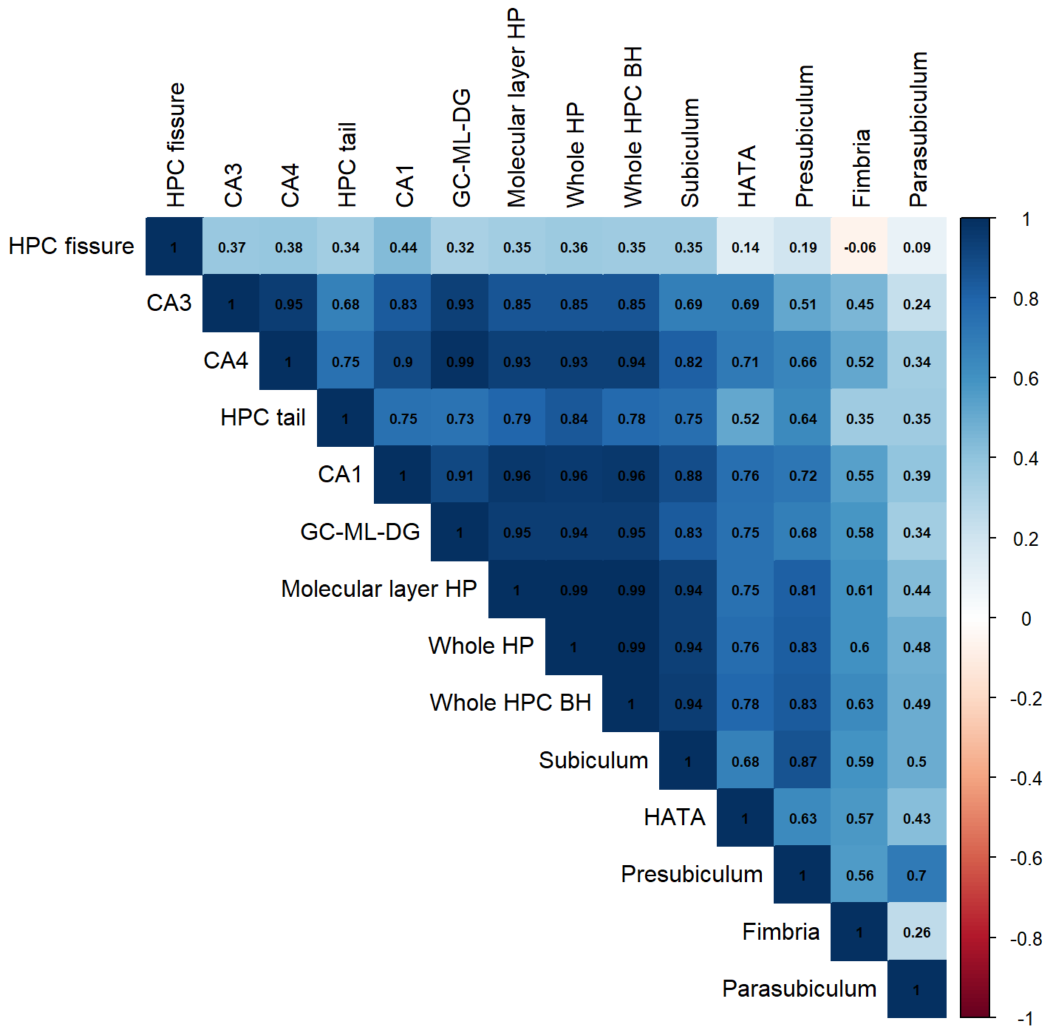


**Supplementary Figure 2**. Hippocampal subregions correlations. HP (Hippocampus), BH (body and head), HPC (Hippocampal), HATA (hippocampus-amygdala-transition-area), GC-ML-DG (Granule Cell and Molecular Layer of the Dentate Gyrus), CA (Cornu ammonis).

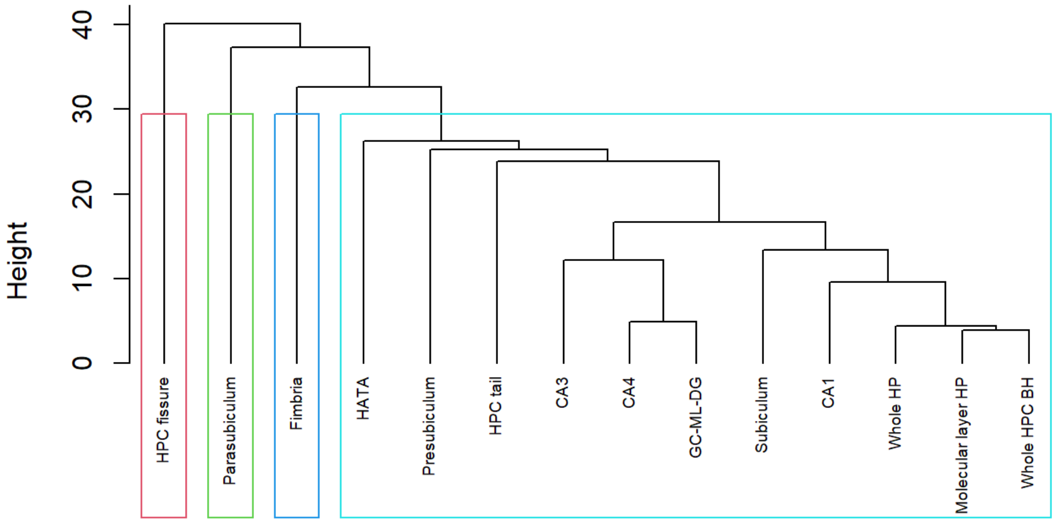


**Supplementary Figure 3**. Cluster dendrogram for hippocampal subfields. HP (Hippocampus), BH (body and head), HPC (Hippocampal), HATA (hippocampus-amygdala-transition-area), GC-ML-DG (Granule Cell and Molecular Layer of the Dentate Gyrus), CA (Cornu ammonis).

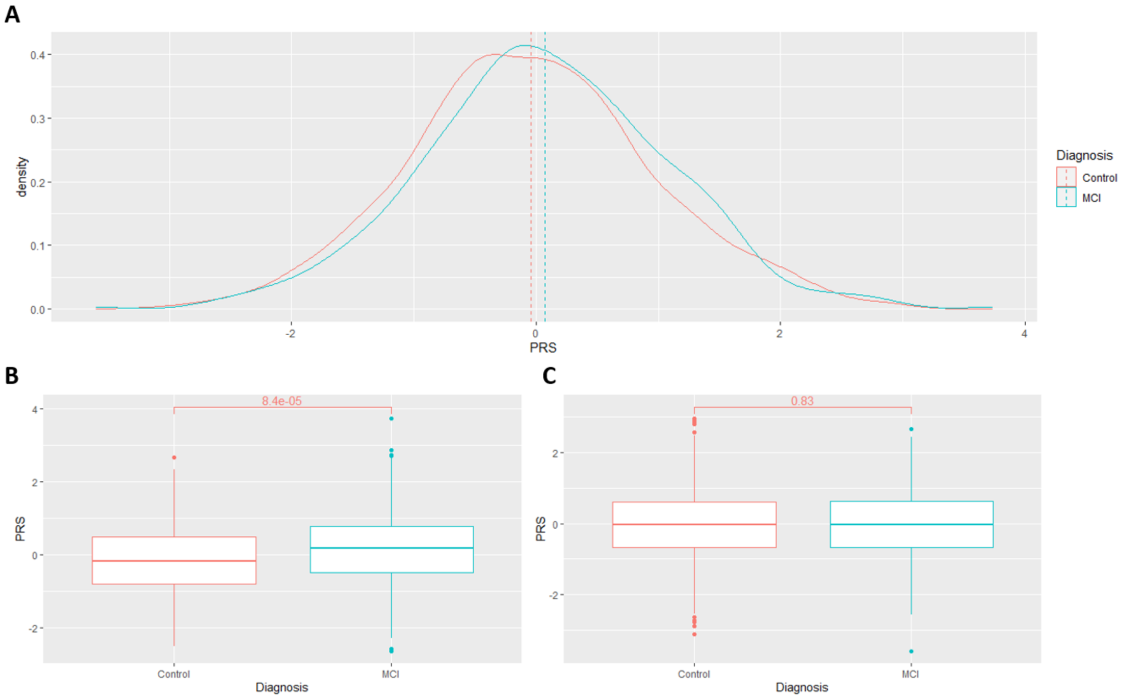


**Supplementary Figure 4**. Distribution of AD-PRS values in mild cognitive impairment (MCI) compared with control individuals (**A**). In *APOE* ε4 carriers (**B**) and *APOE* ε4 non-carriers (**C**).

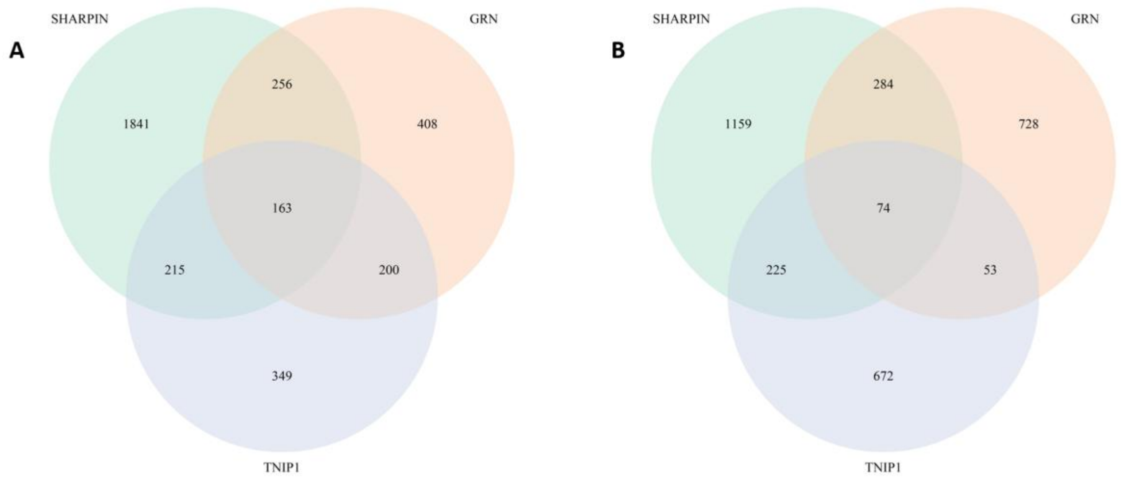


**Supplementary Figure 5**. Venn diagram of the co-expressed genes between *SHARPIN*, *GRN* and *TNIP1* in brain (**A**) and neuron (**B**).
