## Supplementary Methods for "Genetic dissection of hippocampal sclerosis of ageing using magnetic resonance imaging surrogates"

**The Vallecas Project**

The Vallecas Project is a single-centre longitudinal community-based study, currently in its eighth year. Genetic testing was conducted at baseline visit, and yearly clinical, neuropsychological, and MRI assessments remain ongoing. Inclusion and exclusion criteria have been described previously. From a pool of 1213 participants, additional criteria were set, including only those with MRI scanning and without a diagnosis of MCI or dementia, leaving a total of 916 participants on the first visit for cross-sectional analyses, of which 729 had GAWAS data or polymorphisms of interest. All participants provided written informed consent. The Vallecas project was approved by the Ethics Committee of the Instituto de Salud Carlos III.

**ADNI 1**

The ADNI was launched in 2003 as a public-private partnership, led by Principal Investigator Michael W. Weiner, MD. The primary goal of ADNI has been to test whether serial MRI, positron emission tomography (PET), other biological markers, and clinical and neuropsychological assessment can be combined to measure the progression of MCI and early AD.

The first phase of ADNI was designed to find more sensitive and accurate biomarkers for the early detection and tracking of AD. This study was based on thousands of brain scans, genetic profiles, blood and cerebrospinal fluid biomarkers and brain imaging measures including structural MRI and PET from a total of 800 individuals diagnosed with mild cognitive impairment (MCI), early AD and elderly control subjects including 200 elderly control subjects, 400 individuals diagnosed with mild cognitive impairment (MCI) and 200 with early Alzheimer's disease (AD). This study developed improved methods that create uniform standards for acquiring longitudinal, multi-site MRI and PET data on patients with AD, MCI and elderly controls and therefore provides maximum power to determine treatment effects. Moreover, it acquired a generally accessible data repository which describes longitudinal changes in brain structure and metabolism as well as clinical/cognitive and biomarker data for the validation of imaging surrogates.

**ADNI GO**

The ADNI GO study followed the ADNI1 project defining and characterizing the stage of the AD spectrum that precedes MCI by enrolling 200 subjects in the mildest symptomatic phase of AD (EMCI). It includes 500 elderly controls and MCI (rollover from ADNI1), 200 subjects in the mildest symptomatic phase of AD (EMCI) (new for ADNI GO). This study performed F18 amyloid imaging on the CN and LMCI subjects from ADNI1 and the newly enrolled EMCI subjects. FDG PET was performed in association with F18 amyloid imaging. ADNI GO established a national network for F18 amyloid imaging and test hypotheses concerning the prevalence and severity of brain amyloid accumulation and its relationship to current and previous changes of clinical state, MRI, FDG-PET, CSF and plasma biomarkers from ADNI1. Collected 3T MRI on all newly enrolled subjects at Baseline, Month 3, Month 6, and Month 12. Continued longitudinal clinical/cognitive and 1.5T MRI studies of approximately 500 LMCI and Cognitively Normal subjects from ADNI1 for an additional 2 years. Collected and analyzed blood and CSF biomarkers from all newly enrolled EMCI and follow-up subjects. Collected blood samples for DNA and RNA extraction. Newly enrolled subjects will also have samples collected for Cell Immortalization and APOE genotyping.

**ADNI2**

The purpose of ADNI2 was to examine how brain imaging and other biomarkers can be used to measure the progression of mild cognitive impairment (MCI) and early Alzheimer's disease. Together with previous studies ADNI1 and ADNI-GO, ADNI2 seek to determine the relationships among brain imaging results, biomarkers, and clinical, cognitive, and genetic characteristics of the entire spectrum of Alzheimer's as it evolves from normal aging through dementia. The overall goals are increased knowledge concerning the sequence and timing of events leading to MCI and Alzheimer's disease, better methods for early detection of these conditions, and data that informs clinical trials aimed at slowing disease progression. This study includes 150 normal controls (new for ADNI2), 450-500 CN and MCI (rollover from ADNI1), 150 Early Mild Cognitive Impairment (EMCI; new for ADNI2), 200 EMCI (rollover from ADNI GO), 150 Late Mild Cognitive Impairment (LMCI; new for ADNI2) and 200 mild AD (new for ADNI2).

This study determined the relationships among clinical, imaging, genetic, and biochemical biomarker characteristics of the entire spectrum of Alzheimer’s disease (AD) as the pathology evolves. Informed the neuroscience of AD, identified diagnostic and prognostic markers and outcome measures that can be used in clinical trials, and helped develop the most effective clinical trial scenarios. Developed uniform standards for acquiring longitudinal multisite MRI and PET data on patients with AD, MCI, and elderly controls. Performed longitudinal clinical, cognitive, MRI, PET (18F-Florbetapir and FDG), and blood and CSF biomarker studies on 550 newly enrolled subjects in addition to continuing these studies for approximately 700 subjects from ADNI1 and ADNI GO for an additional 5 years. Collected blood samples for DNA and RNA extraction. Newly enrolled subjects will also have samples collected for Cell Immortalization and APOE genotyping. Validated the clinical diagnoses and imaging and biomarker surrogates through neuropathological examination of ADNI1, ADNI GO, and ADNI2 participants who come to autopsy.

**BIOFACE**

BIOFACE is a two-year study of clinical, cognition, and biomarkers in individuals with early-onset mild cognitive impairment (age <65-yo) carried out at Ace Alzheimer Center Barcelona. The study goal is to characterize the different phenotypes from a clinical, neuropsychological, and biomarker point of view and to investigate the cerebrospinal fluid (CSF) and plasma proteomics as well as the role of neuronal-derived plasma exosomes (NPEs) as early biomarkers of AD. Participants underwent extended neurological and neuropsychological batteries, multimodal biomarkers including brain MRI, blood, saliva, CSF, anthropometric, and neuro-ophthalmological examinations.

**EPAD**

The European Prevention of Alzheimer's Dementia (EPAD) is a European collaborative research to expand knowledge about the first stages of Alzheimer's and thus prevent dementia before symptoms appear. EPAD is made up of 38 European institutions, including research centers, universities, European pharmaceutical laboratories and patient associations. The objective of EPAD is to develop an infrastructure that allows carrying out concept tests to accelerate early decision making in the development of candidate drugs and their possible combinations to prevent Alzheimer's disease.

**FACEHBI**

Fundació ACE Healthy Brain Initiative (FACEHBI) is a longitudinal study of biomarkers, risk factors, lifestyle and cognition in a cohort of 200 individuals with Subjective Cognitive Decline (SCD) carried out a Ace Alzheimer Center Barcelona since 2014. SCD refers to the self-perception of cognitive problems, including memory loss, without impairment on standardized cognitive tests *(Jessen et al. 2014, Mitchell et al. 2014).* Participants undergo annual follow-ups with comprehensive neurological exam and cognitive battery. Additionally, at v0, v2, v5 and v8 a longer assessment is performed including self-administered questionnaires, neuroimaging with brain MRI, amyloid positron emission tomography (PET) and retinal optical coherence tomography (OCT) and blood, cerebrospinal fluid and fecal sample collection.

**MOPEAD**

The MOPEAD (Models of Patient Engagement for Alzheimer's Disease) project is a European initiative that aimed to give the citizen an active role in the early detection of Alzheimer's disease. MOPEAD was funded by IMI (Innovative Medicines Initiative) - public-private partnership between the EU and the European Federation of Pharmaceutical Industry Associations (EFPIA). MOPEAD was led by ACE and included the participation of Eli Lilly Company Ltd, ASDM Consulting, and AstraZeneca AB. The European Institute of Women's Health, GMV Soluciones

Globales Internet SAU, Karolinska Institute, KITE Innovation (Europe) Ltd, Spomincica-Alzheimer Slovenia, Cologne University Hospital, Ljubljana University Medical Center, Vall D'Hebron-Institut University Hospital Recerca, Stichting VUmc and Alzheimer Europe. The objective of MOPEAD was to sensitize citizens and increase their participation in the diagnostic process of AD, which also helped in identifying unknown cases. In particular, the project tested the effectiveness of four different models of citizen participation (an online questionnaire, ACE´s Open House Initiative, a test conducted by primary care physicians and a cognitive test for patients diagnosed with type 2 diabetes).

**NTB-Clinic-FRCB-IDIBAPS**

The Neurological Tissue Bank (NTB) at Hospital Clínic-FRCB-IDIBAPS (Fundació de Recerca Clínic Barcelona-Institut d’Investigacions Biomèdiques August Pi i Sunyer) contains a repository of preserved nervous tissue samples that are sourced from deceased patients throughout Catalonia. It is focused on neurodegenerative diseases and currently holds nervous tissue from over 2,300 donors. This study included 332 patients with neuropathological confirmation of Alzheimer's disease.
