## Supplementary Tables for "Genetic dissection of hippocampal sclerosis of ageing using magnetic resonance imaging surrogates"

**Supplementary Table 1**. Impact of covariates in the statistical model for the association of the AD-PRS with hippocampal BH.

| | **Estimate** | **SE** | **p -value** |
|---|---|---|---|
| (Intercept) | 6.8627344 | 1.7681677 | 0.000108 |
| PRS | -0.0842782 | 0.0187226 | 7.18E-06 |
| PC1 | -0.3458448 | 0.6330494 | 0.584916 |
| PC2 | 0.9129954 | 0.6608337 | 0.16727 |
| PC3 | -0.6613513 | 0.6011743 | 0.271434 |
| PC4 | 1.5058116 | 0.635283 | 0.017877 |
| PC5 | -0.8285646 | 0.6340329 | 0.19144 |
| PC6 | -0.7289584 | 0.6260938 | 0.244457 |
| PC7 | -0.6808985 | 0.6319018 | 0.281382 |
| PC8 | -0.2904233 | 0.6324686 | 0.646152 |
| PC9 | 0.5739908 | 0.6231629 | 0.357124 |
| PC10 | 0.2735863 | 0.627776 | 0.663032 |
| Age | -0.0782817 | 0.0478843 | 0.102262 |
| Age2 | 0.0001048 | 0.000328 | 0.749274 |
| Sex | -0.2269309 | 0.0478285 | 2.25E-06 |
| Education | -0.0047679 | 0.0043629 | 0.274616 |
| Diagnosis | -0.7307481 | 0.0575022 | < 2e-16 |
| EstimatedTotalIntraCranialVol | 0.3899171 | 0.0229901 | < 2e-16 |
| CohortADNI2 | -0.1373313 | 0.0581838 | 0.018365 |
| CohortBIOFACE | -0.5668226 | 0.1286176 | 1.11E-05 |
| CohortEPAD | -0.1561474 | 0.1559392 | 0.316798 |
| CohortFACEHBI | -0.8814088 | 0.0904961 | < 2e-16 |
| CohortFCIEN | -0.4404407 | 0.064014 | 8.18E-12 |
| CohortMOPEAD | 0.1346365 | 0.1860851 | 0.469453 |

* AD-PRS (Alzheimer's Disease Polygenic Risk Score), BH (body and head), SE (standard error), PC (Principal Component)

**Supplementary Table 2**. Impact of covariates in the statistical model for the association of the AD-PRS with fimbria.

| | **Estimate** | **SE** | **p-value** |
|---|---|---|---|
| (Intercept) | 7.3080599 | 1.9837879 | 0.000236 |
| PRS | -0.069157 | 0.0210276 | 0.001025 |
| PC1 | -0.9339095 | 0.7104256 | 0.188817 |
| PC2 | 1.1491311 | 0.7419849 | 0.121621 |
| PC3 | 0.1857082 | 0.6737084 | 0.782848 |
| PC4 | -0.1148797 | 0.7129228 | 0.872002 |
| PC5 | 0.5518891 | 0.7116173 | 0.43812 |
| PC6 | -0.0037581 | 0.702996 | 0.995735 |
| PC7 | -0.2322511 | 0.709413 | 0.743414 |
| PC8 | -0.8244041 | 0.7097222 | 0.245555 |
| PC9 | 0.776818 | 0.6993964 | 0.266845 |
| PC10 | -0.4668388 | 0.7047037 | 0.50776 |
| Age | -0.0955454 | 0.0537231 | 0.075492 |
| Age2 | 0.0001592 | 0.000368 | 0.665351 |
| Sex | -0.0340669 | 0.0537976 | 0.526655 |
| Education | -0.0067138 | 0.0048999 | 0.170793 |
| Diagnosis | -0.5389243 | 0.0645266 | < 2e-16 |
| EstimatedTotalIntraCranialVol | 0.1210715 | 0.0258564 | 3.04E-06 |
| CohortADNI2 | -0.197091 | 0.0652525 | 0.002559 |
| CohortBIOFACE | -0.8358005 | 0.1451733 | 1.00E-08 |
| CohortEPAD | -0.2820212 | 0.1749988 | 0.107231 |
| CohortFACEHBI | -0.9464415 | 0.1015777 | < 2e-16 |
| CohortFCIEN | -0.3892324 | 0.0719203 | 7.06E-08 |
| CohortMOPEAD | -0.0097748 | 0.2088322 | 0.962672 |

* AD-PRS (Alzheimer's Disease Polygenic Risk Score), SE (standard error), PC (Principal Component)

**Supplementary Table 3**. Biological processes enriched by loci in cluster A identified with STRING**.**

| term ID | term description | observed gene count | background gene count | strength | false discovery rate |
|---|---|---|---|---|---|
| GO:0048519 | Negative regulation of biological process | 12 | 5313 | 0.53 | 0.0229 |
| GO:0097039 | Protein linear polyubiquitination | 2 | 3 | 3 | 0.0314 |

**Supplementary Table 4**. Biological processes enriched by loci in cluster B identified with STRING.

| term ID | term description | observed gene count | background gene count | strength | false discovery rate |
|---|---|---|---|---|---|
| GO:0001819 | Positive regulation of cytokine production | 7 | 482 | 1.46 | 9.79E-06 |
| GO:0032675 | Regulation of interleukin-6 production | 4 | 151 | 1.72 | 0.0029 |
| GO:0019220 | Regulation of phosphate metabolic process | 7 | 1405 | 0.99 | 0.003 |
| GO:0002684 | Positive regulation of immune system process | 6 | 874 | 1.13 | 0.0031 |
| GO:0009967 | Positive regulation of signal transduction | 7 | 1525 | 0.96 | 0.0031 |
| GO:0031399 | Regulation of protein modification process | 7 | 1560 | 0.95 | 0.0031 |
| GO:0045937 | Positive regulation of phosphate metabolic process | 6 | 912 | 1.11 | 0.0031 |
| GO:0050731 | Positive regulation of peptidyl-tyrosine phosphorylation | 4 | 194 | 1.61 | 0.0031 |
| GO:0051247 | Positive regulation of protein metabolic process | 7 | 1512 | 0.96 | 0.0031 |
| GO:1902996 | Regulation of neurofibrillary tangle assembly | 2 | 3 | 3.12 | 0.0031 |
| GO:0031401 | Positive regulation of protein modification process | 6 | 1018 | 1.06 | 0.0033 |
| GO:0042116 | Macrophage activation | 3 | 62 | 1.98 | 0.0033 |
| GO:0071216 | Cellular response to biotic stimulus | 4 | 232 | 1.53 | 0.0033 |
| GO:0051239 | Regulation of multicellular organismal process | 8 | 2749 | 0.76 | 0.0037 |
| GO:0001932 | Regulation of protein phosphorylation | 6 | 1108 | 1.03 | 0.0038 |
| GO:0051347 | Positive regulation of transferase activity | 5 | 586 | 1.23 | 0.0038 |
| GO:0061518 | Microglial cell proliferation | 2 | 8 | 2.69 | 0.0065 |
| GO:0032496 | Response to lipopolysaccharide | 4 | 314 | 1.4 | 0.0072 |
| GO:0001934 | Positive regulation of protein phosphorylation | 5 | 747 | 1.12 | 0.0083 |
| GO:0002732 | Positive regulation of dendritic cell cytokine production | 2 | 11 | 2.55 | 0.0083 |
| GO:0032755 | Positive regulation of interleukin-6 production | 3 | 98 | 1.78 | 0.0083 |
| GO:0045807 | Positive regulation of endocytosis | 3 | 99 | 1.78 | 0.0083 |
| GO:0051707 | Response to other organism | 6 | 1328 | 0.95 | 0.0083 |
| GO:0043549 | Regulation of kinase activity | 5 | 778 | 1.1 | 0.0086 |
| GO:0002252 | Immune effector process | 4 | 375 | 1.32 | 0.0093 |

| | | | | | |
|---|---|---|---|---|---|
| GO:0010646 | Regulation of cell communication | 8 | 3355 | 0.67 | 0.0093 |
| GO:0023051 | Regulation of signaling | 8 | 3367 | 0.67 | 0.0093 |
| GO:1902947 | Regulation of tau-protein kinase activity | 2 | 13 | 2.48 | 0.0093 |
| GO:1905907 | Negative regulation of amyloid fibril formation | 2 | 13 | 2.48 | 0.0093 |
| GO:0060100 | Positive regulation of phagocytosis, engulfment | 2 | 14 | 2.45 | 0.0097 |
| GO:0009605 | Response to external stimulus | 7 | 2355 | 0.77 | 0.0099 |
| GO:0050776 | Regulation of immune response | 5 | 844 | 1.07 | 0.0099 |
| GO:0150078 | Positive regulation of neuroinflammatory response | 2 | 15 | 2.42 | 0.0099 |
| GO:0010604 | Positive regulation of macromolecule metabolic process | 8 | 3533 | 0.65 | 0.0103 |
| GO:0043254 | Regulation of protein-containing complex assembly | 4 | 413 | 1.28 | 0.0104 |
| GO:0002705 | Positive regulation of leukocyte mediated immunity | 3 | 138 | 1.63 | 0.0112 |
| GO:0031333 | Negative regulation of protein-containing complex assembly | 3 | 142 | 1.62 | 0.0116 |
| GO:0044093 | Positive regulation of molecular function | 6 | 1587 | 0.87 | 0.0116 |
| GO:1902430 | Negative regulation of amyloid-beta formation | 2 | 18 | 2.34 | 0.0116 |
| GO:0002281 | Macrophage activation involved in immune response | 2 | 19 | 2.32 | 0.0118 |
| GO:0030334 | Regulation of cell migration | 5 | 927 | 1.03 | 0.0118 |
| GO:1900221 | Regulation of amyloid-beta clearance | 2 | 19 | 2.32 | 0.0118 |
| GO:0032103 | Positive regulation of response to external stimulus | 4 | 453 | 1.24 | 0.0124 |
| GO:1900225 | Regulation of NLRP3 inflammasome complex assembly | 2 | 20 | 2.29 | 0.0124 |
| GO:0002577 | Regulation of antigen processing and presentation | 2 | 21 | 2.27 | 0.0131 |
| GO:0032680 | Regulation of tumor necrosis factor production | 3 | 164 | 1.56 | 0.0143 |
| GO:1902531 | Regulation of intracellular signal transduction | 6 | 1726 | 0.84 | 0.0143 |
| GO:1902533 | Positive regulation of intracellular signal transduction | 5 | 997 | 0.99 | 0.0143 |
| GO:0033674 | Positive regulation of kinase activity | 4 | 494 | 1.2 | 0.0151 |
| GO:0048583 | Regulation of response to stimulus | 8 | 3931 | 0.6 | 0.0151 |
| GO:0030335 | Positive regulation of cell migration | 4 | 529 | 1.17 | 0.0184 |
| GO:0006622 | Protein targeting to lysosome | 2 | 28 | 2.15 | 0.0187 |

| | | | | | |
|---|---|---|---|---|---|
| GO:0060627 | Regulation of vesicle-mediated transport | 4 | 551 | 1.16 | 0.0211 |
| GO:0001774 | Microglial cell activation | 2 | 31 | 2.1 | 0.0219 |
| GO:0071402 | Cellular response to lipoprotein particle stimulus | 2 | 31 | 2.1 | 0.0219 |
| GO:0071219 | Cellular response to molecule of bacterial origin | 3 | 205 | 1.46 | 0.0229 |
| GO:0045321 | Leukocyte activation | 4 | 574 | 1.14 | 0.023 |
| GO:0045732 | Positive regulation of protein catabolic process | 3 | 214 | 1.44 | 0.0244 |
| GO:0032733 | Positive regulation of interleukin-10 production | 2 | 40 | 1.99 | 0.031 |
| GO:0050850 | Positive regulation of calcium-mediated signaling | 2 | 42 | 1.97 | 0.0324 |
| GO:0032735 | Positive regulation of interleukin-12 production | 2 | 43 | 1.96 | 0.0335 |
| GO:0045859 | Regulation of protein kinase activity | 4 | 663 | 1.07 | 0.034 |
| GO:0043408 | Regulation of MAPK cascade | 4 | 668 | 1.07 | 0.0341 |
| GO:0006955 | Immune response | 5 | 1321 | 0.87 | 0.0366 |
| GO:0051129 | Negative regulation of cellular component organization | 4 | 691 | 1.06 | 0.038 |
| GO:0007165 | Signal transduction | 8 | 4714 | 0.52 | 0.0408 |
| GO:0006952 | Defense response | 5 | 1394 | 0.85 | 0.0447 |
| GO:0048260 | Positive regulation of receptor-mediated endocytosis | 2 | 55 | 1.86 | 0.0484 |
| GO:0048678 | Response to axon injury | 2 | 56 | 1.85 | 0.0497 |

**Supplementary Table 5**. Biological processes enriched by loci in cluster C identified with STRING.

| term ID | term description | observed gene count | background gene count | strength | false discovery rate |
|---|---|---|---|---|---|
| GO:0010813 | Neuropeptide catabolic process | 3 | 4 | 2.42 | 0.0064 |
| GO:0050435 | Amyloid-beta metabolic process | 4 | 18 | 1.89 | 0.0064 |
| GO:0030334 | Regulation of cell migration | 13 | 927 | 0.69 | 0.0087 |
| GO:0002252 | Immune effector process | 8 | 375 | 0.88 | 0.0095 |
| GO:0002684 | Positive regulation of immune system process | 12 | 874 | 0.68 | 0.0095 |
| GO:0006897 | Endocytosis | 9 | 447 | 0.85 | 0.0095 |
| GO:0007166 | Cell surface receptor signaling pathway | 18 | 2040 | 0.49 | 0.0095 |
| GO:0010815 | Bradykinin catabolic process | 3 | 7 | 2.18 | 0.0095 |
| GO:0023052 | Signaling | 30 | 5057 | 0.32 | 0.0095 |
| GO:0030335 | Positive regulation of cell migration | 10 | 529 | 0.82 | 0.0095 |
| GO:0043085 | Positive regulation of catalytic activity | 14 | 1191 | 0.62 | 0.0095 |
| GO:0044093 | Positive regulation of molecular function | 16 | 1587 | 0.55 | 0.0095 |
| GO:0045807 | Positive regulation of endocytosis | 5 | 99 | 1.25 | 0.0095 |
| GO:0048518 | Positive regulation of biological process | 34 | 6207 | 0.28 | 0.0095 |
| GO:0051094 | Positive regulation of developmental process | 14 | 1332 | 0.57 | 0.0095 |
| GO:0051239 | Regulation of multicellular organismal process | 21 | 2749 | 0.43 | 0.0095 |
| GO:0051240 | Positive regulation of multicellular organismal process | 16 | 1505 | 0.57 | 0.0095 |
| GO:0060100 | Positive regulation of phagocytosis, engulfment | 3 | 14 | 1.88 | 0.0095 |
| GO:0060627 | Regulation of vesicle-mediated transport | 10 | 551 | 0.81 | 0.0095 |
| GO:0150094 | Amyloid-beta clearance by cellular catabolic process | 3 | 8 | 2.12 | 0.0095 |
| GO:1902959 | Regulation of aspartic-type endopeptidase activity involved in amyloid precursor protein catabolic process | 3 | 11 | 1.98 | 0.0095 |
| GO:1902991 | Regulation of amyloid precursor protein catabolic process | 4 | 44 | 1.5 | 0.0095 |
| GO:0065009 | Regulation of molecular function | 22 | 3085 | 0.4 | 0.0097 |
| GO:0007165 | Signal transduction | 28 | 4714 | 0.32 | 0.0106 |

| | | | | | |
|---|---|---|---|---|---|
| GO:2000026 | Regulation of multicellular organismal development | 14 | 1389 | 0.55 | 0.0127 |
| GO:1902430 | Negative regulation of amyloid-beta formation | 3 | 18 | 1.77 | 0.0128 |
| GO:0030100 | Regulation of endocytosis | 6 | 209 | 1 | 0.0143 |
| GO:1900221 | Regulation of amyloid-beta clearance | 3 | 19 | 1.74 | 0.0143 |
| GO:0002682 | Regulation of immune system process | 14 | 1438 | 0.53 | 0.0165 |
| GO:0007154 | Cell communication | 29 | 5165 | 0.3 | 0.0171 |
| GO:0051345 | Positive regulation of hydrolase activity | 9 | 589 | 0.73 | 0.0171 |
| GO:1901076 | Positive regulation of engulfment of apoptotic cell | 2 | 2 | 2.55 | 0.018 |
| GO:0010646 | Regulation of cell communication | 22 | 3355 | 0.36 | 0.0233 |
| GO:0043410 | Positive regulation of MAPK cascade | 8 | 481 | 0.77 | 0.0233 |
| GO:0043549 | Regulation of kinase activity | 10 | 778 | 0.66 | 0.0233 |
| GO:0045785 | Positive regulation of cell adhesion | 8 | 485 | 0.76 | 0.0233 |
| GO:0048522 | Positive regulation of cellular process | 30 | 5584 | 0.28 | 0.0233 |
| GO:0050790 | Regulation of catalytic activity | 18 | 2370 | 0.43 | 0.0233 |
| GO:0002274 | Myeloid leukocyte activation | 5 | 150 | 1.07 | 0.0239 |
| GO:0010628 | Positive regulation of gene expression | 12 | 1146 | 0.57 | 0.0239 |
| GO:0010814 | Substance P catabolic process | 2 | 3 | 2.37 | 0.0239 |
| GO:0010875 | Positive regulation of cholesterol efflux | 3 | 26 | 1.61 | 0.0239 |
| GO:0065005 | Protein-lipid complex assembly | 3 | 26 | 1.61 | 0.0239 |
| GO:1902960 | Negative regulation of aspartic-type endopeptidase activity involved in amyloid precursor protein catabolic process | 2 | 3 | 2.37 | 0.0239 |
| GO:0051049 | Regulation of transport | 15 | 1763 | 0.48 | 0.0253 |
| GO:0050865 | Regulation of cell activation | 9 | 658 | 0.68 | 0.0272 |
| GO:0050730 | Regulation of peptidyl-tyrosine phosphorylation | 6 | 259 | 0.91 | 0.0273 |
| GO:0097066 | Response to thyroid hormone | 3 | 29 | 1.56 | 0.0273 |
| GO:0043408 | Regulation of MAPK cascade | 9 | 668 | 0.68 | 0.0285 |
| GO:0045595 | Regulation of cell differentiation | 14 | 1582 | 0.49 | 0.0285 |
| GO:0051716 | Cellular response to stimulus | 32 | 6357 | 0.25 | 0.0285 |

| | | | | | |
|---|---|---|---|---|---|
| GO:0002316 | Follicular B cell differentiation | 2 | 4 | 2.25 | 0.0292 |
| GO:0050776 | Regulation of immune response | 10 | 844 | 0.62 | 0.0292 |
| GO:0050793 | Regulation of developmental process | 18 | 2492 | 0.4 | 0.0292 |
| GO:0050794 | Regulation of cellular process | 45 | 11025 | 0.16 | 0.0292 |
| GO:0051336 | Regulation of hydrolase activity | 11 | 1011 | 0.58 | 0.0292 |
| GO:0051604 | Protein maturation | 6 | 275 | 0.89 | 0.0315 |
| GO:0050867 | Positive regulation of cell activation | 7 | 401 | 0.79 | 0.0326 |
| GO:0019222 | Regulation of metabolic process | 33 | 6784 | 0.23 | 0.0339 |
| GO:1902905 | Positive regulation of supramolecular fiber organization | 5 | 175 | 1 | 0.0339 |
| GO:0038027 | Apolipoprotein A-I-mediated signaling pathway | 2 | 5 | 2.15 | 0.0343 |
| GO:0045917 | Positive regulation of complement activation | 2 | 5 | 2.15 | 0.0343 |
| GO:0051130 | Positive regulation of cellular component organization | 11 | 1049 | 0.57 | 0.0343 |
| GO:0045597 | Positive regulation of cell differentiation | 10 | 876 | 0.6 | 0.0347 |
| GO:0043254 | Regulation of protein-containing complex assembly | 7 | 413 | 0.78 | 0.0355 |
| GO:0032879 | Regulation of localization | 16 | 2103 | 0.43 | 0.036 |
| GO:1902107 | Positive regulation of leukocyte differentiation | 5 | 181 | 0.99 | 0.036 |
| GO:0050777 | Negative regulation of immune response | 5 | 185 | 0.98 | 0.0378 |
| GO:0061097 | Regulation of protein tyrosine kinase activity | 4 | 99 | 1.15 | 0.0383 |
| GO:0010647 | Positive regulation of cell communication | 14 | 1693 | 0.46 | 0.0389 |
| GO:0023051 | Regulation of signaling | 21 | 3367 | 0.34 | 0.0389 |
| GO:0045321 | Leukocyte activation | 8 | 574 | 0.69 | 0.0389 |
| GO:0048523 | Negative regulation of cellular process | 26 | 4736 | 0.29 | 0.0389 |
| GO:0048584 | Positive regulation of response to stimulus | 16 | 2131 | 0.42 | 0.0389 |
| GO:0050789 | Regulation of biological process | 46 | 11655 | 0.14 | 0.0389 |
| GO:0051246 | Regulation of protein metabolic process | 18 | 2622 | 0.38 | 0.0389 |
| GO:1903898 | Negative regulation of PERK-mediated unfolded protein response | 2 | 6 | 2.07 | 0.0389 |
| GO:0031334 | Positive regulation of protein-containing complex assembly | 5 | 193 | 0.96 | 0.0392 |

| | | | | | |
|---|---|---|---|---|---|
| GO:0016192 | Vesicle-mediated transport | 12 | 1298 | 0.51 | 0.0397 |
| GO:0050731 | Positive regulation of peptidyl-tyrosine phosphorylation | 5 | 194 | 0.96 | 0.0397 |
| GO:0051347 | Positive regulation of transferase activity | 8 | 586 | 0.68 | 0.0398 |
| GO:0070050 | Neuron cellular homeostasis | 3 | 41 | 1.41 | 0.0409 |
| GO:0051247 | Positive regulation of protein metabolic process | 13 | 1512 | 0.48 | 0.0413 |
| GO:1902531 | Regulation of intracellular signal transduction | 14 | 1726 | 0.46 | 0.0413 |
| GO:0052547 | Regulation of peptidase activity | 7 | 446 | 0.74 | 0.0422 |
| GO:0002694 | Regulation of leukocyte activation | 8 | 601 | 0.67 | 0.0448 |

**Supplementary Table 6**. Top ten pathways ranked based on FDR threshold enriched by co-expressed genes between *SHARPIN*, *GRN* and *TNIP1* in brain (N = 163) identified by ORA with WebGestalt.

| **Gene Set** | **Description** | **Expect** | **Ratio** | **P-value** | **FDR** |
|---|---|---|---|---|---|
| hsa04141 | Protein processing in endoplasmic reticulum | 1.82 | 7.16 | 2.22 × 10-08 | **7.25 × 10-06** |
| hsa00511 | Other glycan degradation | 0.20 | 20.20 | 3.70 × 10-05 | **6.03 × 10-03** |
| hsa04142 | Lysosome | 1.35 | 5.91 | 5.59 × 10-05 | **6.07 × 10-03** |
| hsa05169 | Epstein-Barr virus infection | 2.21 | 4.07 | 3.34 × 10-04 | **2.73 × 10-02** |
| hsa05163 | Human cytomegalovirus infection | 2.48 | 3.64 | 7.61 × 10-04 | **4.96 × 10-02** |
| hsa04662 | B cell receptor signaling pathway | 0.78 | 6.40 | 1.05 × 10-03 | **5.70 × 10-02** |
| hsa05170 | Human immunodeficiency virus 1 infection | 2.33 | 3.43 | 2.19 × 10-03 | 1.02 × 10-01 |
| hsa05162 | Measles | 1.45 | 4.13 | 3.20 × 10-03 | 1.20 × 10-01 |
| hsa04658 | Th1 and Th2 cell differentiation | 1.01 | 4.94 | 3.31 × 10-03 | 1.20 × 10-01 |
| hsa05131 | Shigellosis | 0.72 | 5.59 | 5.53 × 10-03 | 1.71 × 10-01 |

* Significant FDR (FDR ≤ 0.05) in bold.

† Expect (number of expected genes in the pathway), Ratio (enrichment ratio), FDR (False Discovery Rate).

**Supplementary Table 7.** KEGG pathways significantly enriched by co-expressed genes between *SHARPIN*, *GRN* and *TNIP1* in brain (N = 163) found in the STRING database.

| **Term ID** | **Description** | **Strength** | **FDR** |
|---|---|---|---|
| hsa00511 | Other glycan degradation | 1.43 | 3.00E-03 |
| hsa00600 | Sphingolipid metabolism | 1.02 | 2.60E-02 |
| hsa04141 | Protein processing in endoplasmic reticulum | 0.99 | 6.84E-07 |
| hsa04142 | Lysosome | 0.89 | 2.20E-03 |
| hsa04662 | B cell receptor signaling pathway | 0.89 | 2.45E-02 |
| hsa05162 | Measles | 0.80 | 9.80E-03 |
| hsa05164 | Influenza A | 0.78 | 5.30E-03 |
| hsa05169 | Epstein-Barr virus infection | 0.76 | 3.30E-03 |
| hsa05163 | Human cytomegalovirus infection | 0.65 | 2.45E-02 |
| hsa05131 | Shigellosis | 0.65 | 2.45E-02 |

* Strength (Enrichment Effect = log10(observed / expected)), FDR (False Discovery Rate).

**Supplementary Table 8**. Top ten pathways ranked based on FDR threshold enriched by co-expressed genes between *SHARPIN*, *GRN* and *TNIP1* in neuron (N = 74) identified by ORA with WebGestalt**.**

| **Gene Set** | **Description** | **Expect** | **Ratio** | **P-value** | **FDR** |
|---|---|---|---|---|---|
| hsa00010 | Glycolysis / Gluconeogenesis | 0.36 | 8.44 | 5.22 × 10-03 | 1 |
| hsa04977 | Vitamin digestion and absorption | 0.13 | 15.95 | 6.83 × 10-03 | 1 |
| hsa00052 | Galactose metabolism | 0.16 | 12.35 | 1.12 × 10-02 | 1 |
| hsa00500 | Starch and sucrose metabolism | 0.19 | 10.63 | 1.50 × 10-02 | 1 |
| hsa00520 | Amino sugar and nucleotide sugar metabolism | 0.25 | 7.97 | 2.58 × 10-02 | 1 |
| hsa00524 | Neomycin, kanamycin and gentamicin biosynthesis | 0.03 | 38.27 | 2.59 × 10-02 | 1 |
| hsa00330 | Arginine and proline metabolism | 0.26 | 7.65 | 2.78 × 10-02 | 1 |
| hsa03040 | Spliceosome | 0.70 | 4.32 | 3.17 × 10-02 | 1 |
| hsa04910 | Insulin signaling pathway | 0.72 | 4.19 | 3.42 × 10-02 | 1 |
| hsa00310 | Lysine degradation | 0.31 | 6.49 | 3.42 × 10-02 | 1 |

* Expect (number of expected genes in the pathway), Ratio (enrichment ratio), FDR (False Discovery Rate). Significant FDR (FDR ≤ 0.05) in bold.
